# Prevalence of dental caries and associated risk factors among HIV-positive and HIV-negative adults at an HIV clinic in Kigali, Rwanda

**DOI:** 10.1101/2022.10.04.22280701

**Authors:** Julienne Murererehe, Yolanda Malele-Kolisa, Francois Niragire, Veerasamy Yengopal

## Abstract

**Background:** Dental caries is among the most frequent oral conditions in HIV-positive (HIV+) persons. There is a lack of baseline information on dental caries prevalence and associated risk factors among HIV+ individuals in comparison to HIV-negative (HIV−) people in Rwanda.

**Objective:** This study was conducted to determine the prevalence of dental caries and associated risk factors among HIV+ and HIV-adults at an HIV clinic of Kigali Teaching Hospital (CHUK) in Kigali, Rwanda.

**Methods:** A comparative cross-sectional study was conducted among 200 HIV+ and 200 HIV-adults aged 18 years and above attending the HIV clinic of CHUK. An oral examination was performed by a calibrated examiner. Caries was assessed using the WHO Decayed (D), Missing (M), and Filled Teeth (F) index (DMFT). Descriptive statistics, Chi-square, t-tests, and multiple logistic regression were used to analyze data.

**Results:** Overall, a higher prevalence (50.5%) of HIV+ adults had dental caries experience (DMFT>0) compared to HIV-counterparts (40.5%) (p=0.045). The prevalence of Decayed teeth (D) was also higher (23.5%) among HIV+ participants compared to HIV-persons (13.6%) (p=0.011). The Mean(SD) DMFT scores among HIV+ and HIV-participant were 2.28 (3.68) and 1.29 (2.21) respectively (p=0.001). After performing multiple logistic regression analysis, the predictors of dental caries in HIV+ persons were being a female (OR= 2.33; 95%CI= 1.14-4.75), frequent dental visits (OR= 4.50; 95% CI=1.46-13.86) and detectable RNA viral load (OR= 4.50; 95% CI=1.46-13.86). In HIV-participants, the middle age range (36-45 years), and frequent dental visits were significantly associated with dental caries (OR= 6.61; 95%CI=2.14-20.37) and (OR=3.42; 95%CI: 1.337-8.760) respectively.

**Conclusion:** The prevalence of dental caries was higher in HIV+ adults than in HIV-counterparts. The reported higher prevalence of caries in HIV+ persons was associated with being a female, detectable viral load, and frequent dental visits. Therefore, there is a need for effective oral health interventions specific to HIV+ individuals in Rwanda to raise awareness of the risk of dental caries and provide preventive oral health services among this population. To ensure timely oral health care among HIV+ persons, there is a need for an effort from policymakers and other stakeholders to integrate oral health care services within the HIV treatment program in Rwanda.

## Introduction

Oral health is an integral part of overall health and it is strongly associated with systemic conditions [1]. The prevalence of caries is higher among HIV-positive people (HIV+) than among the general population [2]. Thus, comprehensive and optimal treatment of HIV+ patients requires oral disease management.

Acquired Immune Deficiency Syndrome is considered an important public health problem in developed and developing countries [3]. More than 1.5 million people globally became newly infected with HIV in 2020 [4]. The WHO African region is the most affected region, accounting for more than two-thirds of people living with HIV worldwide [3]. In Rwanda, HIV prevalence is higher in urban areas (7.1%) compared to rural settings (2.3%) [5].

Since the introduction of highly active antiretroviral treatment (HAART) in 1996, the life expectance of HIV-positive (HIV+) people has considerably increased [6,7]. HAART has also significantly reduced the occurrence of HIV-related oral lesions [8]. As HIV+ people live longer, it is imperative to promote their good oral health and access to quality dental care.

Dental care is reported as the least frequently met healthcare need among HIV+ persons [9]. Poor dental function resulting from oral diseases such as dental caries affect the general health of HIV+ individuals [10,11]. Moreover, dental decay negatively affects the physical, mental, and social life of affected people [12]. In addition, oral diseases can affect adherence to antiretroviral therapy (ART) among HIV-positive persons [12]. Also, dental problems among HIV+ individuals are known to be more severe and difficult to manage compared to oral problems among HIV-people[10]. Thus, oral health care should not be ignored and should form an important component of comprehensive treatment available for HIV+ patients.

There is a high burden of oral diseases in Rwanda according to the latest Rwandan National Oral Health Survey. Nearly 65% of participants in the survey had dental caries with more than 54% untreated caries [13]. Data on caries prevalence and associated risk factors among HIV+ individuals is lacking in Rwanda. This study was conducted to provide baseline information on the prevalence of dental caries and associated risk factors among HIV+ adults in comparison to HIV-negative individuals at an HIV clinic of Kigali Teaching hospital (CHUK). The information from this study will enable us to inform policymakers and other stakeholders on the best strategies to prevent oral diseases, specifically dental caries among HIV+ persons thereby contributing to the improvement of their oral health-related quality of life.

## Materials and methods

### Study design and participants

This was a comparative cross-sectional study. The study population consisted of adult HIV– positive (HIV+) and HIV-negative (HIV−) adults attending the HIV clinic of Kigali Teaching Hospital (CHUK). All HIV positive (HIV+) adults, aged 18 years and older, and diagnosed with HIV infection at least 3 months prior to recruitment were included in the study. We also included all HIV-attendees aged 18 years and above who came for HIV voluntary testing and who was diagnosed as HIV−.

### Sample size and sample size calculation

The sample size was 400 participants, including 200 HIV+ and 200 HIV-adults. The sample size for comparative cross-sectional study objectives was calculated using Stata software. Assuming a study power of 80%, considering that the prevalence of caries is 60% in HIV_+_ people and 45% in HIV-persons and considering a ratio of 1[14]. The minimum study sample estimated was 346 participants. Four hundred (400) people were recruited to account for non-respondents.

### Data collection tools and procedures

Participants were informed about the study and recruited at the two sites of the clinic as they attended for care on the recruitment day. All HIV+ participants at the HIV clinic of CHUK were outpatients. The first site of recruitment was at the Voluntary HIV Counselling and Testing (VCT) and it was for the recruitment of HIV-adults. The second site was next to the physicians’ rooms, and it was for recruiting old cases of HIV+ participants. We worked hand in hand with physicians and practitioners who provided HIV results to the patients. To get HIV-persons, respondents were first given their HIV status results by the nurse. After providing the results, the nurse informed people whose HIV results were negative about the ongoing research. HIV-adults who showed interest to participate in the study, were sent to a data collection room to get informed consent. To recruit HIV+ persons, physicians/nurses also informed their patients about the ongoing study. Those who agreed to participate were sent to the researchers for consent signature and data collection. All consenting participants were examined for caries. The Clinical oral examination was performed by a calibrated experienced dental professional working at the University of Rwanda using the WHO (DMFT) index. Participants were seated in a semi-supine position and examined under natural light.

### Statistical analysis

Descriptive statistics including frequencies, percentages and means scores of oral diseases (DMFT mean cores) were computed. The Chi-square test was used to test the relationships between the presence of caries and categorical variables. A t-test was used for data associations between continuous variables. Multiple logistic regression tests were done to determine the relationship of dental caries (dichotomous outcome) with various factors. A separate analysis was done with HIV-related factors (CD4 counts, RNA viral load, type and duration on HIV treatment, WHO staging) as independent variables and adjusted for the sociodemographic, nutritional, and behavioral variables. Kappa score statistical analysis was used to evaluate the intra-examiner reliability during examiner calibration. The level of significance was set at 5%.

### Ethical consideration

The ethical clearances to conduct the study were received from Human Research Ethics Committee (No M200351) from the University of Witwatersrand, Institutional Review Board (No 573/CMHSIRB/2019) from University of Rwanda and Research ethics committee of Kigali Teaching Hospital (No EC/CHUK/026/2020). The informed written consents were given to all participants before data correction. The confidentiality of patients was observed by using an anonymous questionnaire.

## Results

### Demographic characteristics

The demographic characteristics of HIV-positive (HIV+) and HIV-negative (HIV−) adults are presented in table 1. The results revealed a statistically significant difference in age group among HIV+ participants compared to HIV-individuals (p=<0.001). The mean age was significantly higher in HIV+ participants (43.51, 95%CI:41.51-45.51) than the mean age in HIV-persons (36.53, (95% CI: 34.72-38.33) (p<0.001). Also, the majority of HIV+ participants were living in urban areas (81.5%) (n=163) compared to HIV-adults (63%) (p<0.001). More HIV+ participants completed primary (40%) and secondary school (34 %) compared to HIV-individuals who completed primary (37%) and secondary school level (25%) (p=0.020). In addition, significantly (p=0.024) more HIV+ adults were unemployed (32 %) compared to unmployed HIV-persons (22 %). The prevalence of HIV+ persons in ubudehe category 3 and 4 was significantly higher (67.5%) compared to the prevalence of HIV-respondents in ubudehe category 3 and 4 (51,5%) (p=0.005).

**Table 1.**
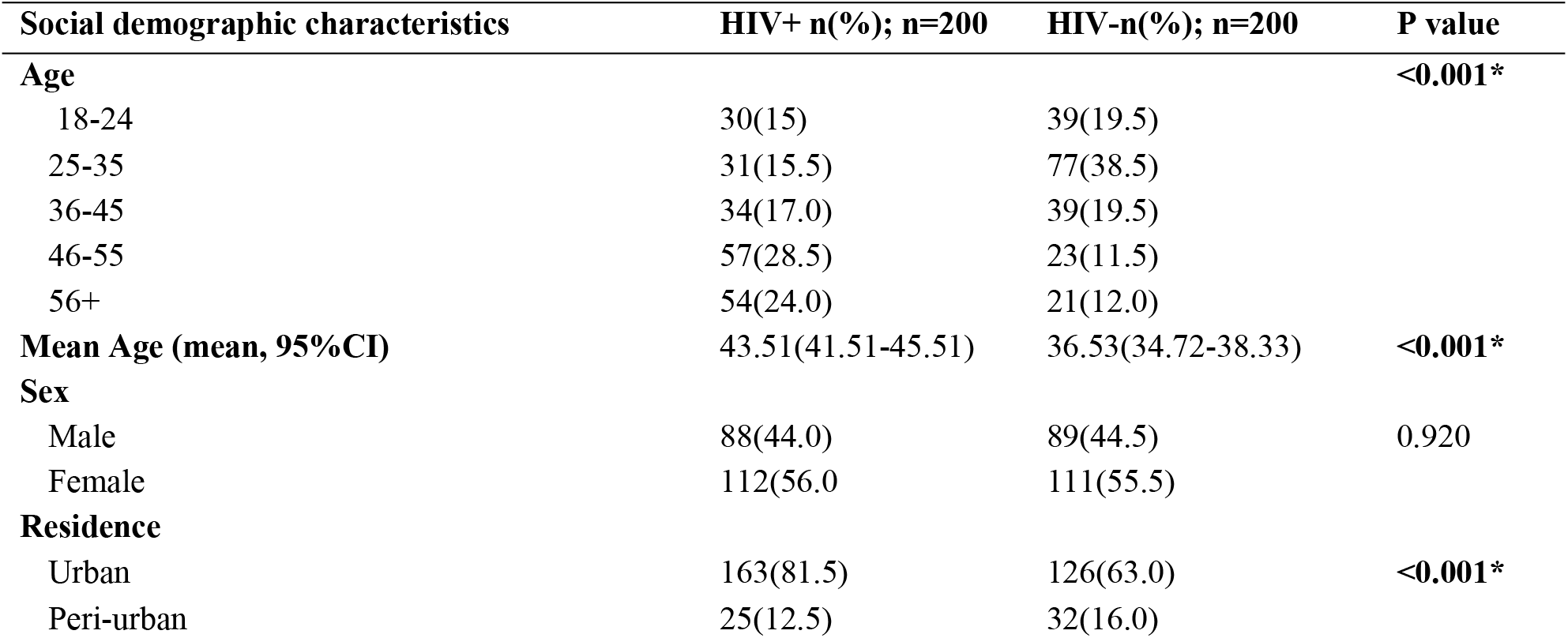
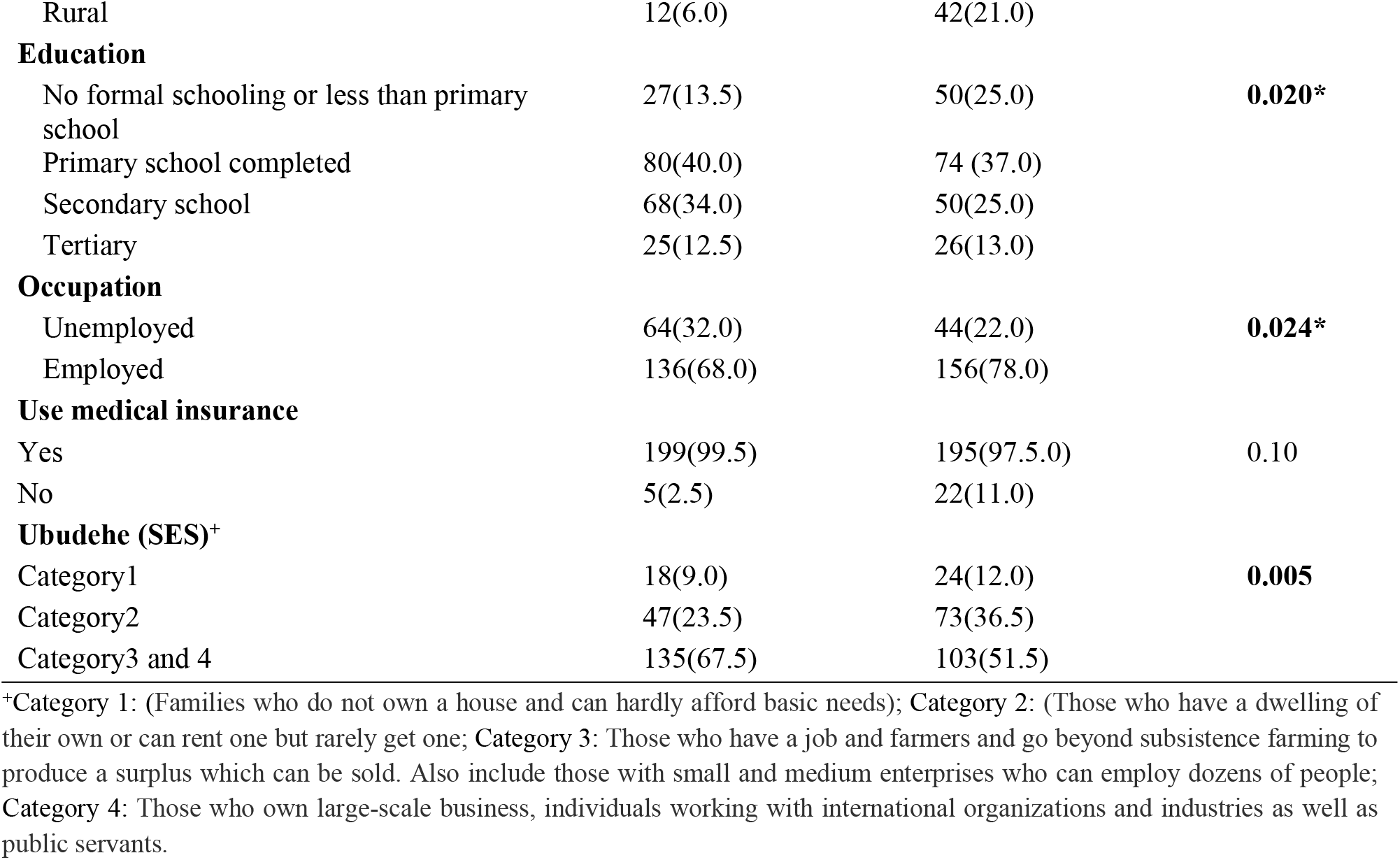
Comparison of Socio demographic characteristics participants.

### Comparison of dental caries among participants according to HIV infection status

The results in Table 2 show that HIV+ participants experienced significantly more caries than HIV-individuals [(Decay (D) 47 (23.5%) vs. 27 (13.6%)]; p< 0.01]. The mean (SD) DMFT scores for HIV+ and HIV-persons were 2.28 (3.68) and 1.29 (2.21) respectively, (p=**0.001)**.

**Table 2.** Distribution of dental caries according to HIV infection status.

| Variable | HIV+ (n=200) |  | HIV-(n=200) |  | P-value |
| --- | --- | --- | --- | --- | --- |
|  | n(%)Yes | n(%) No | Yes N(%) | n(%) No |  |
| D | 47(23.5) | 153(76.5) | **27(13.6)** | 172(86.4) | **0.011\*** |
| M | 80(40) | 120(60) | 69(34.5) | 131(65.5) | 0.255 |
| F | 23(11.6) | 176(88.4) | 14(7) | 186(93) | 0.117 |
| DMFT | 101(50.5) | 99(49.5) | 81(40.5) | 119(59.5) | **0.045\*** |
|  | **Mean** | **SD** | **Mean** | **SD** | **P value** |
| D | 0.57 | 1.42 | 0.17 | 0.47 | **<0.001\*** |
| M | 1.36 | 2.78 | **0.96** | 1.84 | 0.091 |
| F | 0.37 | 1.31 | **0.17** | 0.78 | 0.069 |
| DMFT | 2.28 | 3.68 | 1.29 | 2.21 | **0.001\*** |

### Comparison of factors associated with dental caries among HIV+ and HIV-adults

The results in table 3 showed that HIV+ females were 2.33 times more likely than HIV+ males to have dental caries (95% CI= 1.142-4.252). Although not significant, HIV-females were also 1.37 times more likely than HIV-males to have dental caries 1.37 (95%CI= 0.680-2.774).

**Table 3.**
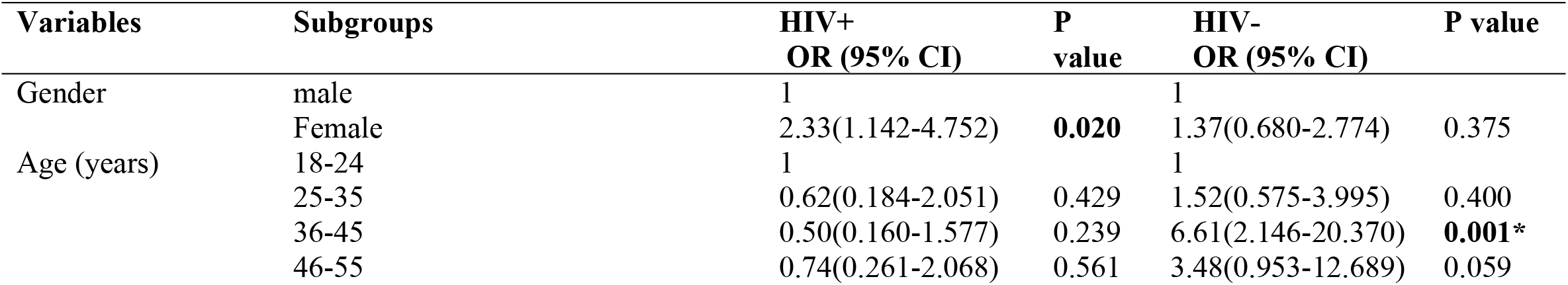
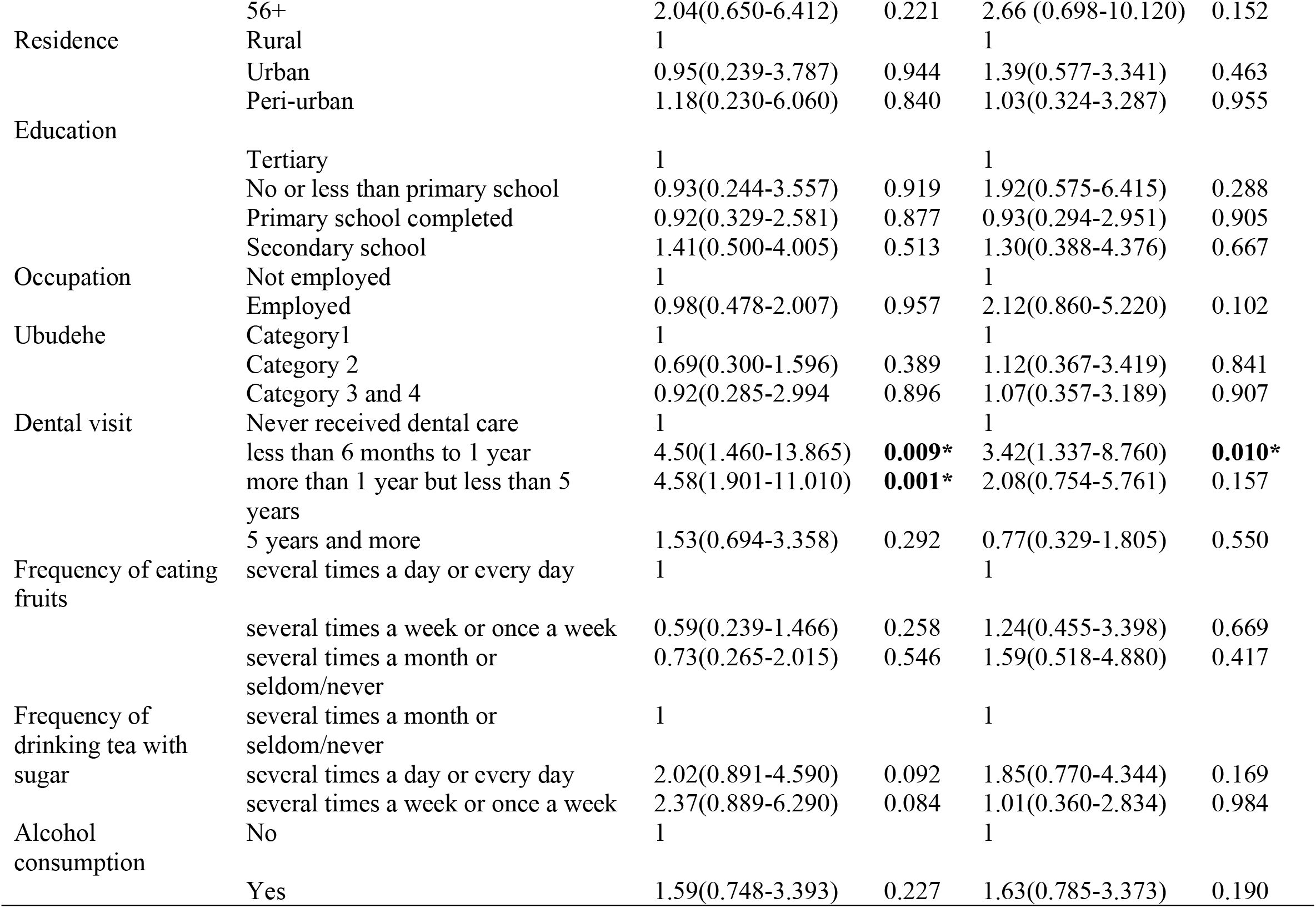
Comparison of factors associated with dental caries among participants according to HIV infection.

**Table 4^+^.** Logistic regression analysis of the HIV-related factors associated with caries among HIV-positive adults.

| Variable | Subgroups | OR ( 95% CI) | P value |
| --- | --- | --- | --- |
| CD4 | Group I ($\geq 500$) | 1 | |
|  | Group II (200-499) | 0.904(0.440-1.856) | 0.784 |
|  | Group III (<200) | 2.150(0.572-8.075) | 0.257 |
| RNA-Viral Load | Undetectable | 1 |  |
|  | Detectable | 2.691(1.004-7.208) | **0.049\*** |
| WHO staging | Stage I | 1 |  |
|  | Stage II To IV | 2.130(0.658-6.895) | 0.207 |
| Types of ART | classII and III | 1 |  |
|  | Class I | 1.707(0.703-4.143) | 0.237 |
| Duration on ARVs | 11-15 years | 1 |  |
|  | 1-5 years | 1.089(0.313-3.788) | 0.893 |
|  | 6-10 years | 0.988(0.436-2.237) | 0.978 |
| Duration of HIV-infection | 1-8 years | 1 |  |
|  | 9-16 years | 0.566(0.1552.059) | 0.388 |
|  | 17+ years | 1.176(0.281-4.909) | 0.824 |

\*Adjusted for gender, age, residence, education, occupation, ubudehe (SES), dental visit, frequency of eating fruits, frequency of drinking tea with sugar and alcohol consumption.

Amongst HIV-adults, participants aged 36-45 years were 6.61 times more likely than the younger age group (18-14) to develop dental caries (95% CI=2.146-20.370), the result was statistically significant (p=0.001). The remaining age categories were not statistically significant.

HIV-respondents who have visited a dentist within less than 6 months to 1-year period were 3.24 times more likely to have dental caries (95%CI= 1.34-8.76) compared to HIV-adults who never visited dentists. The difference was statistically significant. Participants who visited dentists after 1 year but less than 5 years were approximately twice more likely to have dental caries (95%CI=0.75-5.76) than the research participants who never received dental care. Those who visited dentist after 5 years were 1.30 times less likely to have dental caries (95% CI=0.329-1.805) than those who never visited a dentist.

### Multiple logistic regression analysis of HIV-related factors associated with dental caries among HIV+ adults

After adjusting for all factors not related to HIV infection (gender, age, residence, education, occupation, ubudehe (income status), dental visit, frequency of eating fruits, frequency of drinking tea with sugar and alcohol consumption), the results revealed that HIV+ participants with detectable RNA-Viral Load (≥20) were 2.69 times more likely (95% CI= 1.004-7.208) to have dental caries than participants with undetectable RNA-Viral Load and the results were statically significant (p =0.049).

## Discussion

The findings of this study revealed a significantly higher prevalence of dental caries in HIV+ participants compared to HIV-individuals. Amongst HIV-positive (HIV+) respondents, cohort females, frequent dental visits, and detectable RNA-viral load were significantly associated with dental caries. In HIV-participants, caries prevalence was associated with frequent dental visits and the age range of 36-45. The findings of this study are in line with growing evidence of a higher prevalence and risk of dental caries among HIV+ people compared to the general population [15–17].

The higher prevalence of caries in HIV+ adults than in HIV-individuals is similar to results by other scholars in developed and developing countries [16,18–20]. Untreated dental caries or decay (D) was also more prevalent in HIV+ participants 47(23.5%) than in HIV-individuals 27(13.6%). This may be explained by the unavailability of oral health education and caries preventive services within HIVclinics and services throughout the country.

AlthoughRwanda has shown remarkable improvement in HIV services and HIV+ can easily access ARVs, counseling services, regular checkups for non-communicable and other infectious diseases [21], no oral health services available at these specialized sevices. In addition, there are no regular prevention services, no program to raise awareness about dental caries and to inform HIV+ persons about dental caries risks and effect in existing HIV services or at the community level. This may contribute to a higher prevalence of dental caries among HIV+ individuals than HIV-persons. In their research, Feng and colleagues emphasized that oral health programs that target HIV+ perons should consider affordable dental care, a stigma-free setting, care delivered safely, and an accessible locations [22]. Thus, the need for an oral health program that can benefit HIV+ people in Rwanda.

In contrast to the present study, the results of research by Malele-Kolisa and colleagues, in South Africa, younger adolescents living with HIV had reported lower caries prevalence (D) than those of undiagnosed schools children [23]. This may be explained by contextual factors where there were oral health strategies and programs that targeted HIV+ adolescents and allowed them to have timely access to regular oral health education and preventive services in South Africa.

The Rwandan first National oral health strategic plan has highlighted barriers that may be associated with the burden of oral diseases in the country. The major part of oral health services is provided in District and referral hospitals. Very few primary health posts and centers can provide dental services. Many private dental clinics are mostly located in the City of Kigali and some other secondary cities. Primary oral health service is only provided in the form of analgesics for pain release in health posts and health centers and all patients are referred to hospitals for oral health care [24]

Since oral healthcare is provided in hospitals, it is not easy for the majority of Rwandans (who normally access primary health care at health posts and health centers level) to access oral health care due to the long distance to the hospitals. These issues result in the abandonment of oral health care for many patients. In addition, there is an overload of patients in hospitals with a shortage of oral health providers [24]

Moreover, in Rwanda District hospitals, there is a shortage of infrastructure including the lack of adequate space and functional dental equipment such as dental chairs to provide oral health services [24]. Apart from the recent and very first National strategic plan, there is no specific policy or plan that is specifically dedicated to oral health in Rwanda. Oral health is integrated into different policy and regulatory documents related to None Communicable Diseases (NCDs) and there is no focal person in charge of oral health at the Ministry of health level to ensure the coordination and address issues related to oral health care in the country [24]. All those problems related to oral health services in Rwanda are not exceptional to HIV+ individuals and since this group is more exposed to oral health problems including dental caries, it is very important to develop oral health interventions specific to HIV+ people in Rwanda.

In our study, dental caries was significantly associated with being a female among HIV+ participants. These findings are similar to recent results of a study done in Uganda by Kalanzi and colleagues where HIV+ females had a higher prevalence of dental caries than HIV+ males [25]. Although not significant, HIV-females were also more affected by dental caries than HIV-males. The literature highlights different factors that expose women to dental caries than males in the general population. Women’s family role has been shown as one of the factors for the higher prevalence of dental caries compared to males. Commonly, women have been family member with the responsibility for food preparation. This allows them to have access to foods and snacks between meals more frequently than males, which increases their chance of caries development compared to men [26,27].

In addition, women go through hormonal changes during puberty, menstruation, and pregnancy and this can modify the saliva flow making the oral environment more prone to caries for women than for men [26,27]. For that reason, there is a need to develop vivacious oral health strategies that will target women in particular, especially in high-risk groups such as HIV-positive persons.

Based on their exposure, women, especially those who are HIV+, have to be given greater attention when planning preventive and oral health education interventions to decrease the risk of dental caries among this group. To increase the number of women who access dental services, previous interventions have considered home visits and oral health programs to provide oral education, and preventive oral health services [28]. Therefore, to increase accessibility to preventive oral health care among women HIV+, there is a need for contextual oral health models among this population in Rwanda.

The results of our study revealed that a higher prevalence of HIV + and HIV-adults who frequently visited dentists experienced more caries compared to participants who less frequently visited dentists. Consistent results were reported in the literature [29,30]. As highlighted by other researchers, the DMFT index takes into account the past treatment history. Therefore, the DMFT index is affected by extracted teeth (M) and the fillings done due to dental caries (F). In our study, the M (Missing teeth due to caries) was the most prevalent component of DMFT in the group of HIV+ and HIV-people. Having a higher prevalence of people with M component suggests that participants in our study visited dentists at a late stage of dental caries development where it was not possible to save their teeth. It also implies that our study population never visited dentists for a check-up and preventive dental services or these services simply did not exist in the study area where patients were recruited. Thus, the need to raise awareness of the benefit of regular and timely dental visits among HIV+ and HIV-individuals in Rwanda.

On the other hand, the higher prevalence of caries experience among participants who frequently visited dentists may suggest their poor oral healthcare-seeking behavior. The issue of poor oral health-seeking behavior was previously discussed in the literature, especially among people in Sub-saharan Africa where oral diseases are overlooked by the general population and people only visit dentists in cases of emergencies and for symptomatic reasons [31].

Amongst HIV-adults, middle age (36-45 years) was significantly associated with dental caries experience. A study from four countries (England and Wales, the United States, Japan, and Sweden) revealed a larger increase in DMFT in adulthood compared to the young group [32]. A recent study done in Ethiopia has also highlighted that age increase is significantly associated with caries experience [33]. However, some researchers had reported contrasting results where young age was associated with increased dental caries compared to adulthood [34]. These findings may be explained by various factors accross different community. For example, in many of developing countries, factors such as increase of sugary diet in young people, inavailability of preventive services, lack of awareness on how to do good oral hygiene and high cost of dental treatments expose young adults to caries than adults [33].

The results of our study also revealed a significant association between detectable HIV RNA-viral load and caries among HIV+ respondents after adjusting for the other factors not related to HIV-infection. Similar to the results of our study, different researchers have also found an association between HIV RNA-viral load with dental caries [6,23,35,36]. These findings may suggest that dental caries can affect the immunity of HIV+ individuals. However, based on the design of our results, it is not possible to determine the causal relationship between dental caries and higher RNA viral load among HIV+ persons. Thus, further studies are needed to give more clarifications on the mechanism of this relationship. On the other hand, HIV+ persons with immune suppression are reported to experience a reduction of saliva flow (xerostomia) [16]. The decreased saliva flaw contributes to easy development of dental caries due to lack saliva buffering capacity that controls the PH of mouth environment [37]. Based on the relationship of RNA viral load and dental caries, there is a need for oral health strategies to provide oral health education and early preventive dental services among HIV+ people in Rwanda.

## Strength of the study

In Rwanda, this is the first study to look at the prevalence of dental caries and associated risk factors among HIV+ persons in comparison to HIV-individuals. It is also among the few studies that compared HIV+ and HIV-adults in regards to caries and its associated risk factors in Sub-saharan Africa.

## Limitations

The results of the study may not be generalizable to the general population of Rwanda because the study was done in an urban HIV clinic in Kigali city which could not represent the whole country. In addition, a cross-sectional study design makes it difficult to establish causality. Large longitudinal studies are needed to better understand the nature of the associations between caries and various risk factors.

## Conclusion

Overall, the prevalence of dental caries was higher in HIV+ than in HIV-participants. The reported higher prevalence of caries in HIV+ persons was associated with being female, detectable viral load, and frequent dental visits. Therefore, there is a need for effective oral health interventions specific to HIV+ individuals in Rwanda to raise awareness of the risk of dental caries and provide preventive oral health services among this population. To ensure timely oral health care among HIV+ persons, there is a need for effort from policymakers and stakeholders to integrate oral health care services within HIV treatment programs in Rwanda. We furthermore recommend further prospective studies that will analyze in depth the strength of the association of caries with different factors among HIV+ persons in comparison to HIV-individuals.

## Data Availability

All relevant data are within the manuscript and its Supporting Information files.

## Acknowledgment

We Thank Dr Babatunde Adedokun from the university of Ibadan for providing his expertise that greatly assisted data analysis, interpretation, and discussion of findings.

## Author Contributions

**Conceptualization:** Julienne Murererehe, Yolanda Malele Kolisa, Veerasamy Yengopal.

**Data curation:** Julienne Murererehe, Yolanda Malele Kolisa, Veerasamy Yengopal

**Formal analysis:** Julienne Murererehe, Francois Niragire, Yolanda Malele Kolisa, Veerasamy Yengopal.

**Funding acquisition:** Julienne Murererehe

**Investigation:** Julienne Murererehe, Yolanda Malele Kolisa, Veerasamy Yengopal

**Methodology:** Julienne Murererehe, Yolanda Malele Kolisa, Francois Niragire, Veerasamy Yengopal

**Supervision:** Yolanda Malele Kolisa, Veerasamy Yengopal

**Writing_ original draft:** Julienne Murererehe, Yolanda Malele Kolisa, Francois Niragire, Veerasamy Yengopal.

**Writing_ review & editing:** Julienne Murererehe, Yolanda Malele Kolisa, Francois Niragire, Veerasamy Yengopal.

## Supporting information

**S1 file** Dataset “DTA”

## Notes

### Competing Interest Statement

The authors have declared no competing interest.

### Clinical Trial

N/A

### Funding Statement

JM was funded by the Consortium for Advanced Research Training in Africa (CARTA). CARTA is jointly led by the African Population and Health Research Center and the University of the Witwatersrand and funded by the Carnegie Corporation of New York (Grant No. G-19-57145), Sida (Grant No:54100113), Uppsala Monitoring Center, Norwegian Agency for Development Cooperation (Norad), and by the Wellcome Trust [reference no. 107768/Z/15/Z] and the UK Foreign, Commonwealth & Development Office, with support from the Developing Excellence in Leadership, Training, and Science in Africa (DELTAS Africa) programme. The statements made and views expressed are solely the responsibility of the Fellow. The funders had no role in study design, data collection, and data analysis, decision to publish, or preparation of the manuscript. URL: www.cartafrica.org

## References

1. Sabbah W, Folayan MO, El Tantawi M. The Link between Oral and General Health. Int J Dent [Internet]. 2019 May 29;2019:1–2. Available from: https://www.hindawi.com/journals/ijd/2019/7862923/

2. Brondani MA, Phillips JC, Kerston RP, Moniri NR. Stigma Around Hiv in Dental Care: Patients’ Experiences. J Can Dent Assoc (Tor) [Internet]. 2016;82:g1–g1. Available from: http://search.ebscohost.com/login.aspx?direct=true&db=ccm&AN=113676385&site=ehost-live

3. WHO. THE GLOBAL HEALTH OBSERVATORY. 2021.

4. WHO. Global HIV Epidemic. 2020.

5. Ministry of Health. NATIONAL HIV/AIDS TARGETS 2018-2020-2030. 2015;1–29.

6. Kikuchi K, Yasuoka J, Tuot S, Okawa S, Yem S, Chhoun P, et al. Dental caries in association with viral load in children living with HIV in Phnom Penh, Cambodia : a cross - sectional study. BMC Oral Health. 2021;159(21):1–8.

7. Weiss RA. Special Anniversary Review : Twenty-five years of human immunodeficiency virus research : successes and challenges. Br Soc Immunol Clin Exp Immunol. 2008;152:201–10.

8. Nicolatou-Galitis O, Velegraki A, Paikos S, Economopoulou P, Stefaniotis T, Papanikolaou IS. HIV Disease / Oral Medicine Effect of PI-HAART on the prevalence of oral lesions in HIV-1 infected patients. A Greek study. Oral Dis. 2004;10(July 2003):145–50.

9. Adedigba MA, Adekanmbi T, Asa S. Pattern of Utilisation of Dental Health Care Among HIV-positive Adult Nigerians. 2016;14(3):215–25.

10. Brondani MA, Phillips JC, Kerston RP, Moniri NR. Stigma Around Hiv in Dental Care: Patients’ Experiences. J Can Dent Assoc (Tor) [Internet]. 2016;(December):g1–g1. Available from: http://search.ebscohost.com/login.aspx?direct=true&db=ccm&AN=113676385&site=ehost-live

11. Yengopal V, Naidoo S. Do oral lesions associated with HIV affect quality of life? Oral Surgery, Oral Med Oral Pathol Oral Radiol Endodontology. 2008;106(1):298–309.

12. Dios PD, Scully C. Antiretroviral therapy : effects on orofacial health and health care. Oral Dis. 2014;20(January 2013):136–45.

13. Morgan JP, Isyagi M, Ntaganira J, Gatarayiha A, E. Pagni S, Roomian TC, et al. Building oral health research infrastructure: the first national oral health survey of Rwanda. Glob Health Action [Internet]. 2018;11(1):298–309. Available from: https://doi.org/10.1080/16549716.2018.1477249

14. Ashagrie M. Which formula to use to calculate the sample size for a cross sectional comparative study? 2020.

15. Nugraha AP, Mensana MP, Soebadi B, Husada D, Astha E, Prasetyo RA, et al. Correlation of Low CD4 + Counts with High Dental Caries Prevalence in Children Living with Perinatal HIV / AIDS Undergoing Antiretroviral Therapy. 2019;1–7.

16. Palupi R, Sosiawan A, Gilang R, Ramadhani A. Factors Influencing Dental Caries in HIV / AIDS Patients. Acta Med Philipp. 2019;53(5):412–6.

17. Rajonson N, Meless D, Ba B, Faye M, Diby JS, N’zore S, et al. High prevalence of dental caries among HIV-infected children in West Africa compared to uninfected siblings. J Public Health Dent. 2017;77(3):234–43.

18. Kumar S, Mishra P, Warhekar S, Airen B, Jain D, Godha S. Oral Health Status and Oromucosal Lesions in Patients Living with HIV/AIDS in India: A Comparative Study. AIDS Res Treat [Internet]. 2014;2014:480247. Available from: http://www.ncbi.nlm.nih.gov/entrez/query.fcgi?cmd=Retrieve&db=PubMed&dopt=Citation&list_uids=25215229

19. Cavasin Filho JC, Giovani EM. Xerostomy, dental caries and periodontal disease in HIV+ patients. Braz J Infect Dis [Internet]. 2009;13:13–7. Available from: http://www.ncbi.nlm.nih.gov/pubmed/19578624

20. Birungi N, Fadnes LT, Marie I, Stadskleiv E, Tumwine JK, Lie SA, et al. Caries experience by socio-behavioural characteristics in HIV-1-infected and uninfected Ugandan mothers – a multilevel analysis. Acta Odontol Scand [Internet]. 2022;80(2):91–8. Available from: https://doi.org/10.1080/00016357.2021.1942544

21. Nsanzimana S, Kanters S, Remera E, Forrest JI, Binagwaho A, Condo J, et al. HIV care continuum in Rwanda: A cross-sectional analysis of the national programme. Lancet HIV [Internet]. 2015;2(5):e208–15. Available from: http://dx.doi.org/10.1016/S2352-3018(15)00024-7

22. Feng I, Brondani M, Bedos C, Donnelly L. Access to oral health care for people living with HIV / AIDS attending a community-based program. Can J Dent Hyg. 2020;54(1):7–15.

23. Kolisa YM, Yengopal V, Shumba K, Igumbor J. The burden of oral conditions among adolescents living with HIV at a clinic in Johannesburg, South Africa. PLoS One. 2019;14(10):1–14.

24. MinistryofHealth. National Oral Health trategic Plan 2019-2024. 2019.

25. Kalanzi D, Mayanja-kizza H, Nakanjako D, Mwesigwa CL, Ssenyonga R, Amaechi BT. Prevalence and factors associated with dental caries in patients attending an HIV care clinic in Uganda : a cross sectional study. 2019;1–8.

26. Ferraro M, Vieira AR. Explaining Gender Differences in Caries : A Multifactorial Approach to a Multifactorial Disease. 2 Int J Dent. 2010;1–5.

27. Kessler JL. A Literature Review on Women’s Oral Health Across the Life Span. Nurs Womens Health. 2017;21(2):108–21.

28. Lam SC, Traylor DO, Anderson EE. Primary Care Recommendations for Oral Health Care in HIV + Patients [Internet]. 2014. Available from: https://www.centerfororalhealth.org/wp-content/uploads/2018/11/Oral-Health-Policy-HIV-2014-6-27-14.pdf

29. Costa SM, Vasconcelos M, Haddad JPA, Henrique M, Abreu NG. The severity of dental caries in adults aged 35 to 44 years residing in the metropolitan area of a large city in Brazil : a cross-sectional study. BMC Oral Health. 2012;12(25).

30. Varenne B, Paris F, Petersen PE, Geneva S, Ouattara S. Oral health behaviour of children and adults in urban and rural areas of Burkina Faso, Africa. Int Dent J. 2006;56(2):61–70.

31. Varenne B, Petersen PE, Fournet F, Msellati P, Gary J, Ouattara S, et al. Illness-related behaviour and utilization of oral health services among adult city-dwellers in Burkina Faso: Evidence from a household survey. BMC Health Serv Res. 2006;6:1–11.

32. Bernabé E, Sheiham A. Age, Period and Cohort Trends in Caries of Permanent Teeth in Four Developed Countries. Am J Public Health [Internet]. 2014 Jul;104(7):e115–21. Available from: https://ajph.aphapublications.org/doi/full/10.2105/AJPH.2014.301869

33. Bogale B, Engida F, Hanlon C, Prince MJ, Gallagher JE. Dental caries experience and associated factors in adults: a cross-sectional community survey within Ethiopia. BMC Public Health [Internet]. 2021 Dec 21;21(1):180. Available from: https://bmcpublichealth.biomedcentral.com/articles/10.1186/s12889-021-10199-9

34. Oscarson N, Espelid I, Jönsson B. Is caries equally distributed in adults? A population-based cross-sectional study in Norway – the TOHNN-study. Acta Odontol Scand [Internet]. 2017 Nov 17;75(8):557–63. Available from: https://doi.org/10.1080/00016357.2017.1357080

35. Souza AJ de, Gomes-Filho IS, Silva CAL da, Passos-Soares J de S, Cruz SS da, Trindade SC, et al. Factors associated with dental caries, periodontitis and intra-oral lesions in individuals with HIV / AIDS*. AIDS Care - Psychol Socio-Medical Asp AIDS/HIV. 2017;0121:1–8.

36. Chen F, Cheng Y, Xie T. Oral Health Status of Young People Infected with HIV in High Epidemic Area of China. J Multidiscip Healthc ©. 2021;14:831–7.

37. Malekipour MR, Messripour M, Shirani F. Buffering capacity of saliva in patients with active dental caries. Asian J Biochem. 2008;3(5):280–3.

